# Multicenter Evaluation of Myocardial Flow Reserve as a Prognostic Marker for Mortality in ¹³N-Ammonia PET Myocardial Perfusion Imaging

**DOI:** 10.1101/2025.06.24.25330229

**Authors:** Giselle Ramirez, Valerie Builoff, Robert JH Miller, Mark Lemley, Isabel Carvajal-Juarez, Erick Alexanderson, Thomas L. Rosamond, Na Song, Mark I. Travin, Leandro Slipczuk, Andrew J. Einstein, Samuel Wopperer, Marcelo Di Carli, Panithaya Chareonthaitawee, Piotr Slomka

**Affiliations:** Departments of Medicine (Division of Artificial Intelligence in Medicine), Imaging and Biomedical Sciences, Cedars-Sinai Medical Center, Los Angeles, CA, USA; Department of Cardiac Sciences, University of Calgary, Calgary AB, Canada; Department of Nuclear Cardiology, Ignacio Chavez National Institute of Cardiology, Mexico City, Mexico; Faculty of Medicine, National Autonomous University of Mexico, Mexico City, Mexico; Department of Cardiovascular Medicine, The University of Kansas Medical Center, Kansas City, KS, USA; Department of Radiology (Nuclear Medicine), Montefiore Medical Center and Albert Einstein College of Medicine, Bronx, NY, USA; Cardiology Division, Montefiore Health System/Albert Einstein College of Medicine, New York, NY, USA; Division of Cardiology, Department of Medicine, and Department of Radiology, Columbia University Irving Medical Center and New York-Presbyterian Hospital, New York, New York, United States; Department of Cardiovascular Medicine, Mayo Clinic, Rochester, MN, USA; Division of Nuclear Medicine and Molecular Imaging, Department of Radiology, Brigham and Women’s Hospital, Boston, MA, USA

**Keywords:** Myocardial flow reserve, ^13^N-ammonia, Myocardial perfusion imaging, All-cause mortality

## Abstract

**Background:** Myocardial flow reserve (MFR), measured by PET MPI, provides valuable information on epicardial coronary disease, diffuse atherosclerosis, and microvascular function. Despite its routine use, the prognostic efficacy of ^13^N-ammonia PET MFR remains unconfirmed in larger multicenter cohorts of patients with suspected or known coronary artery disease (CAD).

**Methods:** We considered patients from five sites in the REFINE PET registry who underwent ^13^N-ammonia PET MPI for CAD. Clinical and imaging data were collected at the time of MPI. MFR was quantified as the ratio of stress to rest myocardial blood flow, using QPET software (Cedars-Sinai Medical Center, Los Angeles, CA). The primary outcome was all-cause mortality (ACM). Survival analyses were performed using Kaplan-Meier and Cox regression models adjusted for clinical and imaging covariates.

**Results:** In total, 6277 patients were included (mean age of 64 years, 56% male). Median follow-up time was 3.8 years. There were 1895 patients with MFR ≤2 and 4382 with MFR >2. Patients with MFR ≤2 had significantly higher mortality than those with MFR >2 (n=701 [37.0%] vs. n=537 [12.3%], respectively; p<0.001). Annualized ACM rates by MFR and SSS ranged from 1.7 to 11.6. In multivariable analysis, MFR ≤2 was independently associated with increased ACM in the overall population (HR 2.70, 95% CI 2.41-3.03, p<0.001), even among patients with no perfusion defects (HR 2.36, 95% CI 1.93-2.89; p<0.001). Mortality risk decreased across increasing MFR deciles ranging from HR 2.73 (95% CI 2.39-3.11) to HR 0.35 (95% CI 0.25-0.49).

**Conclusion:** In this large multicenter cohort, MFR derived from ^13^N-ammonia PET MPI is a strong, independent predictor of ACM, even in patients with normal perfusion. An MFR of ≤2.0 identifies elevated risk, while higher values are associated with improved survival. These findings support the routine integration of MFR to enhance risk stratification in patients with suspected or known CAD.

## INTRODUCTION

Coronary artery disease (CAD) remains the leading cause of morbidity and mortality worldwide (1). As such, accurate risk stratification is essential to guide clinical decision-making and improve patient outcomes. Myocardial flow reserve (MFR), quantified non-invasively using positron emission tomography (PET) myocardial perfusion imaging (MPI), integrates the effects of epicardial coronary stenosis, diffuse atherosclerosis, and microvascular dysfunction (2, 3). Normal MFR suggests preserved coronary physiology, while reduced MFR may reflect impairment in one or more of these domains. Numerous studies with mostly Rb82, have shown that reduced MFR is independently associated with higher rates of adverse outcomes, irrespective of patient demographics, socioeconomic status, or cardiovascular risk factors (3-5). While conventional PET-derived relative perfusion measures provide important insight into flow-limiting epicardial stenoses, they may underestimate diffuse atherosclerosis or microvascular dysfunction that can also contribute to adverse outcomes (3). As a result, MFR has emerged as a valuable tool for enhancing risk stratification in patients with known or suspected CAD.

MFR, particularly when derived from PET MPI using ^13^N-ammonia, is FDA-approved and widely utilized in clinical practice as a diagnostic tool (6). ^13^N-ammonia may show higher quality images, due to its shorter positron range (2.5 mm vs. 8.1 mm for Rb82), and high extraction fraction (7). However, its prognostic utility has not been comprehensively evaluated to date in a large, multicenter cohort using commercially available clinical software for PET MPI quantification. Despite growing recognition of MFR’s physiologic value, its integration into routine decision-making has been limited by the lack of outcome-based data using high-resolution PET tracers including ¹³N-ammonia.

To address these gaps, we assessed the relationship between MFR measured by ^13^N-ammonia PET and all-cause mortality (ACM) in a large, multicenter patient population with suspected or established CAD. This study aimed to determine whether MFR provides independent and incremental prognostic value beyond traditional cardiovascular risk factors and relative perfusion abnormalities.

## METHODS

### Study Population

We retrospectively analyzed 6520 consecutive patients from five sites in the REgistry of Fast Myocardial Perfusion Imaging with NExt generation PET (REFINE PET) who had suspected or established CAD and underwent ^13^N-ammonia PET MPI from. Patients with missing stress total perfusion deficit (n=22), rest total perfusion deficit (n=15), MFR (n=60), prior cardiac transplant (n=155), or treadmill tested (n=19) were excluded. Since some patients met more than one exclusion criterion, a total of 243 unique patients were excluded. In total, 6277 patients were included in the analysis: Brigham and Women’s Hospital (n=2245), University of Kansas Medical Center (n=589), Mayo Clinic (n=1966), National Autonomous University of Mexico (n=437) and Montefiore Medical Center (n=1040). This study was approved by the institutional review boards at each participating site. Approval of the overall study was provided by the Cedars-Sinai Medical Center institutional review board. The study complied with the Declaration of Helsinki, and sites either obtained written informed consent or a waiver of consent for the use of the deidentified data, and sites.

### Clinical Variables

Clinical information was collected at the time of imaging and included age, sex, body mass index (BMI), family history of CAD, current smoking status, previous myocardial infarction (MI), prior revascularization, and history of hypertension, diabetes, dyslipidemia, and heart failure. Each site collected this information using site-specific procedures. Prior CAD was defined as a history of MI or revascularization. The frequency of missing demographic or clinical variables is summarized in **Supplementary Table 1.**

### PET protocol

Patients underwent rest and stress PET MPI following established clinical guidelines (8). All patients fasted for at least three hours, and a 6-minute rest list-mode acquisition was initiated immediately before the administration of ^13^N-ammonia. Pharmacological stress was induced using regadenoson (n=5134, 81.8%), adenosine (n=1012, 16.1%), dobutamine (n=79, 1.26%), or dipyridamole (n=52, 0.83%). A 6-minute stress imaging acquisition was initiated simultaneously with the tracer injection. At peak stress, an additional dose of tracer was administered, and stress images were obtained using the same protocol. A low-dose helical CT scan was performed before both rest and stress PET acquisitions for attenuation correction, as previously described (9). Imaging data were electronically transferred from the five sites to the core laboratory where trained technologists blinded to clinical and outcome information reprocessed and reconstructed the images using specialized software (QPET, Cedars-Sinai Medical Center, Los Angeles, California).

### Myocardial Perfusion Analysis

PET image quantification was performed automatically in batch mode using specialized, previously validated and commercially available software (QPET, Cedars-Sinai Medical Center, Los Angeles, California). Rest and stress relative perfusion were quantified using total perfusion deficit (TPD) (10, 11). Normal perfusion was defined as summed stress score (SSS) < 4 (12). The extent of reversible perfusion defects was determined by the summed difference score (SDS) (13, 14).

### MBF and MFR quantification

Automatic motion correction for dynamic PET-MPI was applied. Absolute MBF (ml/min/g) was quantified at rest and stress using parametric polar maps and measured using a 2-compartment model for ^13^N-ammonia PET (15). MBF and the spillover fraction from the blood to the myocardium were determined via numeric optimization. Stress and rest flow values were computed for each sample on the polar map. Global MFR was calculated as the ratio of stress to rest MBF for the entire left ventricle. Abnormal MFR was defined as ≤2 (15).

### Clinical Scoring

Experienced physicians at each site visually evaluated PET/CT scans during clinical reporting, with access to all available data, including stress and rest TPD, gated functional data, MBF, MFR, other quantitative metrics, CT images, and clinical information. The final visual assessment was based on SSS and summed rest scores (SRS) using the 17-segment American Heart Association model (16). The SDS was calculated as the difference between SSS and SRS and was used as the final clinical score. If visual scores were not available, quantitative SSS and SDS were used. An SSS ≥4 considered to indicate presence of perfusion abnormality (12). SDS ≥2 was considered to indicate a reversible perfusion defect, and SDS <2 was considered to indicate a fixed or non-reversible defect (13, 14).

### Outcome

The primary outcome was ACM, which was determined using the National Death Index for sites in the USA and through patient or family follow-up in Mexico. The secondary outcome was the classification of cardiovascular versus non-cardiovascular death, which was determined from electronic medical records at each site.

### Statistical Analysis

Categorical variables are presented as n (%) and continuous variables are presented as mean (±standard deviation [SD]) if normally distributed or median (interquartile range [IQR]) if not normally distributed. Categorical variables were compared with using a Pearson’s Chi-squared test and continuous variables were compared using a Wilcoxon rank sum test or Kruskal-Wallis test when applicable. Survival analyses for ACM were plotted using the Kaplan-Meier estimator. The unadjusted Kaplan-Meier curves were annotated with an adjusted hazard ratio from the Cox model, which accounts for age and sex and includes a 95% confidence interval (CI). Univariate and multivariate Cox regression models were used to assess the association between MFR and ACM with the time of ^13^N-ammonia PET as the point of entry. Multivariate Model 1 was adjusted for MFR, age, sex, and prior CAD (defined as history of myocardial infarction or revascularization). Multivariate Model 2 was additionally adjusted for BMI, heart rate, and LVEF. Hazard ratios (HRs) are presented along with 95% CI. Survival distributions between MFR groups were compared using the log-rank test.

Annualized mortality rates were calculated by dividing the total events (either ACM or cardiovascular deaths) over the total person years and multiplied by 100. Decile bar charts were generated by dividing MFR into ten equally divided groups. Each decile group was adjusted for sex and age and compared to the rest of the population. The continuous adjusted hazard ratio was adjusted for sex and age as a continuous variable expressed as a cubic spline with three knots. Imputation was made for missing clinical or demographical information used for multivariate Cox regression models. Mode imputation was used for categorical variables, while median imputation was used for continuous variables. The variables with missing values are listed in **Supplementary Table 1**. p<0.05 was considered statistically significant. All analyses were performed using R studio (R version 4.4.1 Inc, Boston, MA).

### Independent Data Access and Analysis

GR and PS had full access to all the data in the study and takes responsibility for its integrity and the data analysis.

## RESULTS

### Patient Characteristics

**Table 1** summarizes clinical and demographic characteristics of the patients categorized by MFR. The study included a total of 6277 patients, with a median age of 65 (IQR 56, 73) and 56% male. The median MFR was 2.5 (1.9-3.2) and 1895 (30.2%) patients had an abnormal MFR (≤2). Patients with MFR ≤2 tended to be older and had a higher prevalence of cardiovascular risk factors and cardiac disease. Nearly all ^13^N-ammonia PET variables demonstrated significant differences between patients with MFR ≤2 and those with MFR >2 (**Table 1**).

**Table 1:**
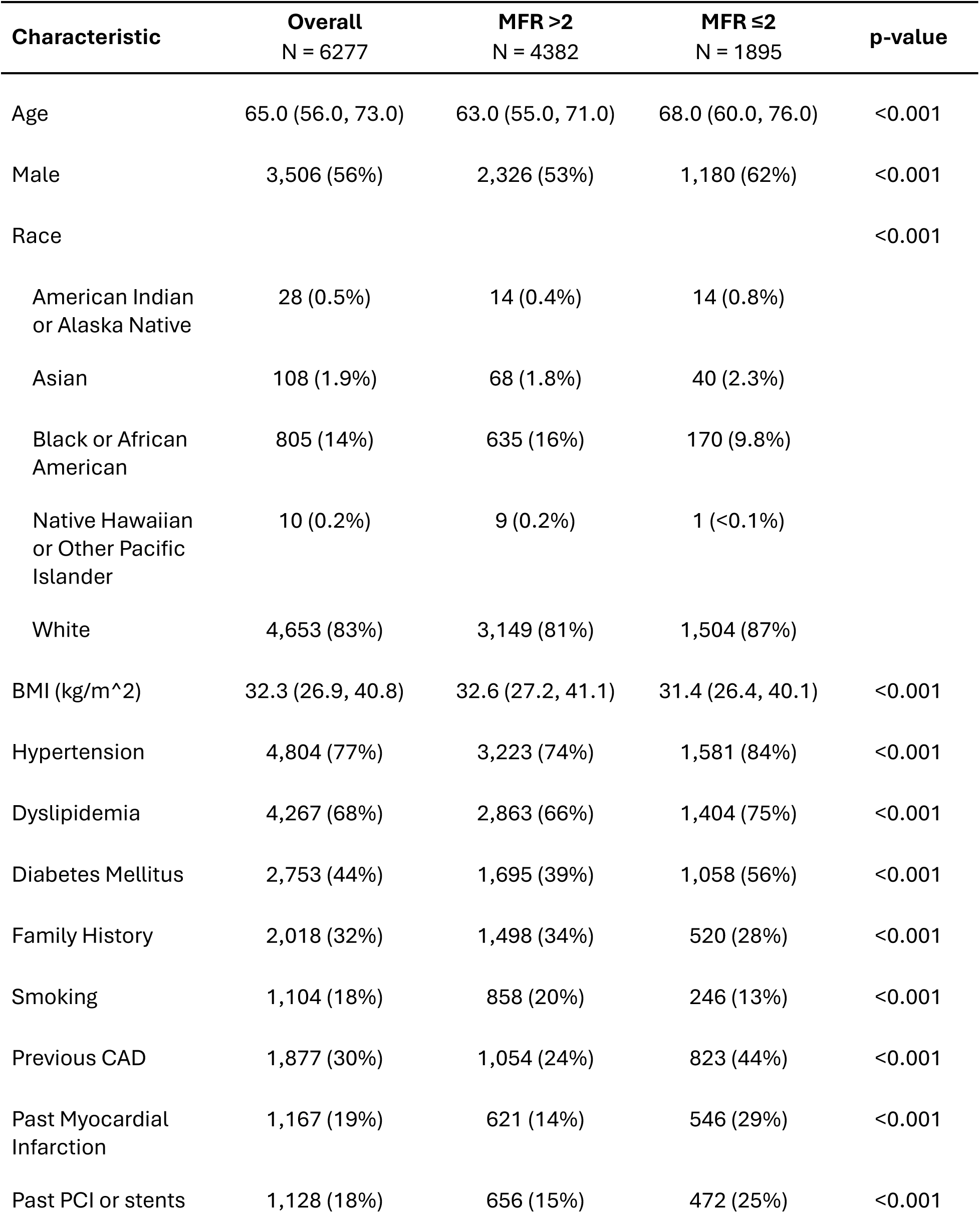
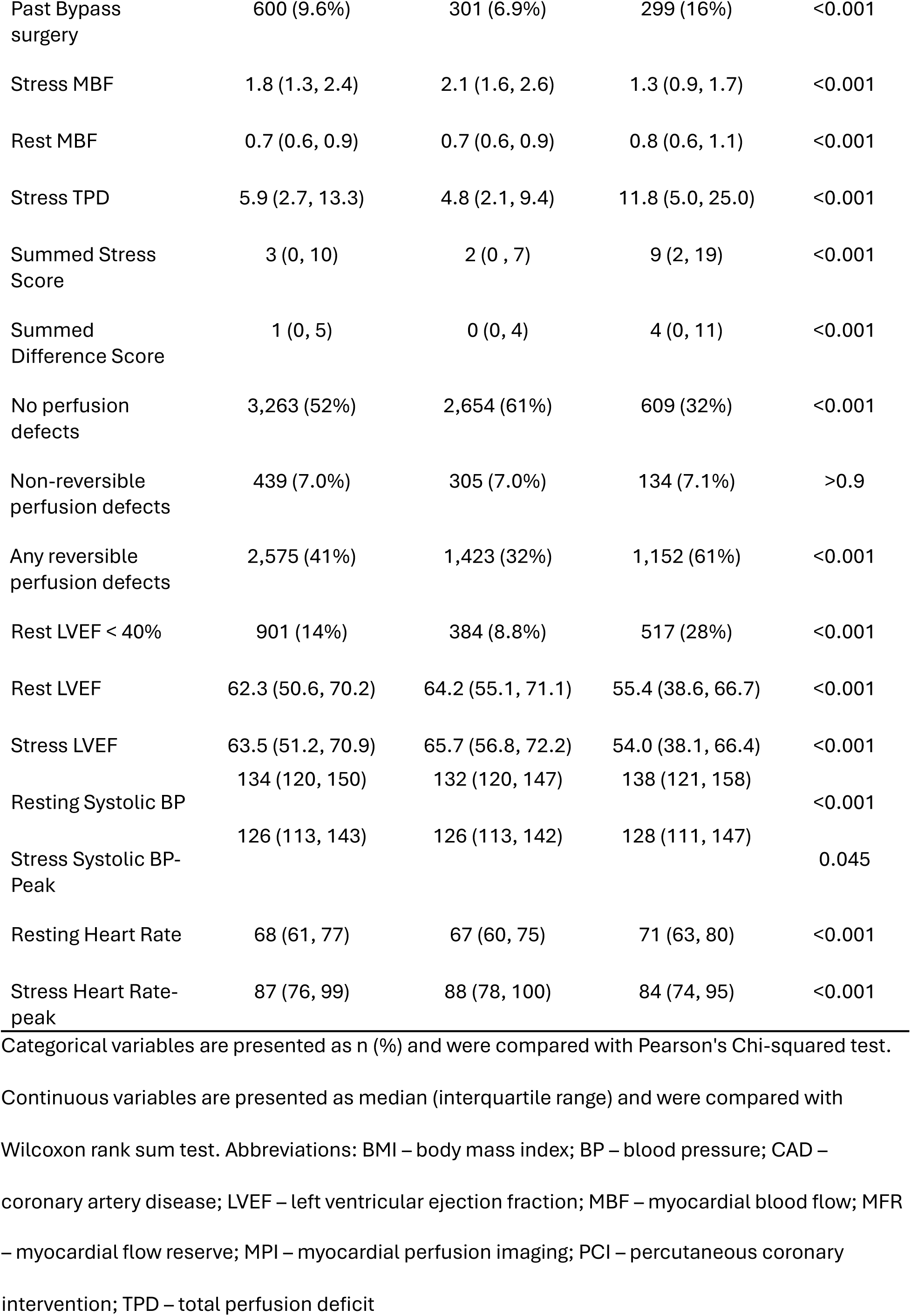
Baseline Characteristics Stratified by MFR.

| Characteristic | Overall<br>N = 6277 | MFR >2<br>N = 4382 | MFR ≤2<br>N = 1895 | p-value |
| --- | --- | --- | --- | --- |
| Age | 65.0 (56.0, 73.0) | 63.0 (55.0, 71.0) | 68.0 (60.0, 76.0) | <0.001 |
| Male | 3,506 (56%) | 2,326 (53%) | 1,180 (62%) | <0.001 |
| Race |  |  |  | <0.001 |
| American Indian<br>or Alaska Native | 28 (0.5%) | 14 (0.4%) | 14 (0.8%) |  |
| Asian | 108 (1.9%) | 68 (1.8%) | 40 (2.3%) |  |
| Black or African<br>American | 805 (14%) | 635 (16%) | 170 (9.8%) |  |
| Native Hawaiian<br>or Other Pacific<br>Islander | 10 (0.2%) | 9 (0.2%) | 1 (<0.1%) |  |
| White | 4,653 (83%) | 3,149 (81%) | 1,504 (87%) |  |
| BMI (kg/m^2) | 32.3 (26.9, 40.8) | 32.6 (27.2, 41.1) | 31.4 (26.4, 40.1) | <0.001 |
| Hypertension | 4,804 (77%) | 3,223 (74%) | 1,581 (84%) | <0.001 |
| Dyslipidemia | 4,267 (68%) | 2,863 (66%) | 1,404 (75%) | <0.001 |
| Diabetes Mellitus | 2,753 (44%) | 1,695 (39%) | 1,058 (56%) | <0.001 |
| Family History | 2,018 (32%) | 1,498 (34%) | 520 (28%) | <0.001 |
| Smoking | 1,104 (18%) | 858 (20%) | 246 (13%) | <0.001 |
| Previous CAD | 1,877 (30%) | 1,054 (24%) | 823 (44%) | <0.001 |
| Past Myocardial<br>Infarction | 1,167 (19%) | 621 (14%) | 546 (29%) | <0.001 |
| Past PCI or stents | 1,128 (18%) | 656 (15%) | 472 (25%) | <0.001 |
| Past Bypass surgery | 600 (9.6%) | 301 (6.9%) | 299 (16%) | <0.001 |
| Stress MBF | 1.8 (1.3, 2.4) | 2.1 (1.6, 2.6) | 1.3 (0.9, 1.7) | <0.001 |
| Rest MBF | 0.7 (0.6, 0.9) | 0.7 (0.6, 0.9) | 0.8 (0.6, 1.1) | <0.001 |
| Stress TPD | 5.9 (2.7, 13.3) | 4.8 (2.1, 9.4) | 11.8 (5.0, 25.0) | <0.001 |
| Summed Stress Score | 3 (0, 10) | 2 (0, 7) | 9 (2, 19) | <0.001 |
| Summed Difference Score | 1 (0, 5) | 0 (0, 4) | 4 (0, 11) | <0.001 |
| No perfusion defects | 3,263 (52%) | 2,654 (61%) | 609 (32%) | <0.001 |
| Non-reversible perfusion defects | 439 (7.0%) | 305 (7.0%) | 134 (7.1%) | >0.9 |
| Any reversible perfusion defects | 2,575 (41%) | 1,423 (32%) | 1,152 (61%) | <0.001 |
| Rest LVEF < 40% | 901 (14%) | 384 (8.8%) | 517 (28%) | <0.001 |
| Rest LVEF | 62.3 (50.6, 70.2) | 64.2 (55.1, 71.1) | 55.4 (38.6, 66.7) | <0.001 |
| Stress LVEF | 63.5 (51.2, 70.9) | 65.7 (56.8, 72.2) | 54.0 (38.1, 66.4) | <0.001 |
| Resting Systolic BP | 134 (120, 150) | 132 (120, 147) | 138 (121, 158) | <0.001 |
| Stress Systolic BP-Peak | 126 (113, 143) | 126 (113, 142) | 128 (111, 147) | 0.045 |
| Resting Heart Rate | 68 (61, 77) | 67 (60, 75) | 71 (63, 80) | <0.001 |
| Stress Heart Rate-peak | 87 (76, 99) | 88 (78, 100) | 84 (74, 95) | <0.001 |

Categorical variables are presented as n (%) and were compared with Pearson's Chi-squared test.
Continuous variables are presented as median (interquartile range) and were compared with
Wilcoxon rank sum test. Abbreviations: BMI – body mass index; BP – blood pressure; CAD –
coronary artery disease; LVEF – left ventricular ejection fraction; MBF – myocardial blood flow; MFR
– myocardial flow reserve; MPI – myocardial perfusion imaging; PCI – percutaneous coronary intervention; TPD – total perfusion deficit

There was a higher frequency of no perfusion defects in participants with an MFR >2 (2654 [61%]) compared to those with MFR ≤2 (609 [32%]; p<0.001). Meanwhile, patients with impaired MFR were more likely to have reversible perfusion defects than patients with preserved MFR (1423 [32%] for MFR >2 vs. 1152 [61%] for MFR ≤2; p<0.001), whereas there was no statistical significance between patients with impaired or preserved for non-reversible perfusion defects (305 [7%] for MFR >2 vs. 134 [7.1%] for MFR ≤2; p>0.9. Additionally, patients with MFR ≤2 demonstrated higher resting MBF (median, 0.8 [IQR, 0.6, 1.1]) and lower stress MBF (1.3 [0.9, 1.7]) compared to those with MFR >2 (0.7 [0.6, 0.9] and 2.1 [1.6, 2.6], respectively; p<0.001 for all). A stratified analysis of ¹³N-ammonia PET findings by extent of reversible and non-reversible perfusion defects, demonstrating a statistically significant difference of p<0.001 is detailed in **Table 2**.

**Table 2.**
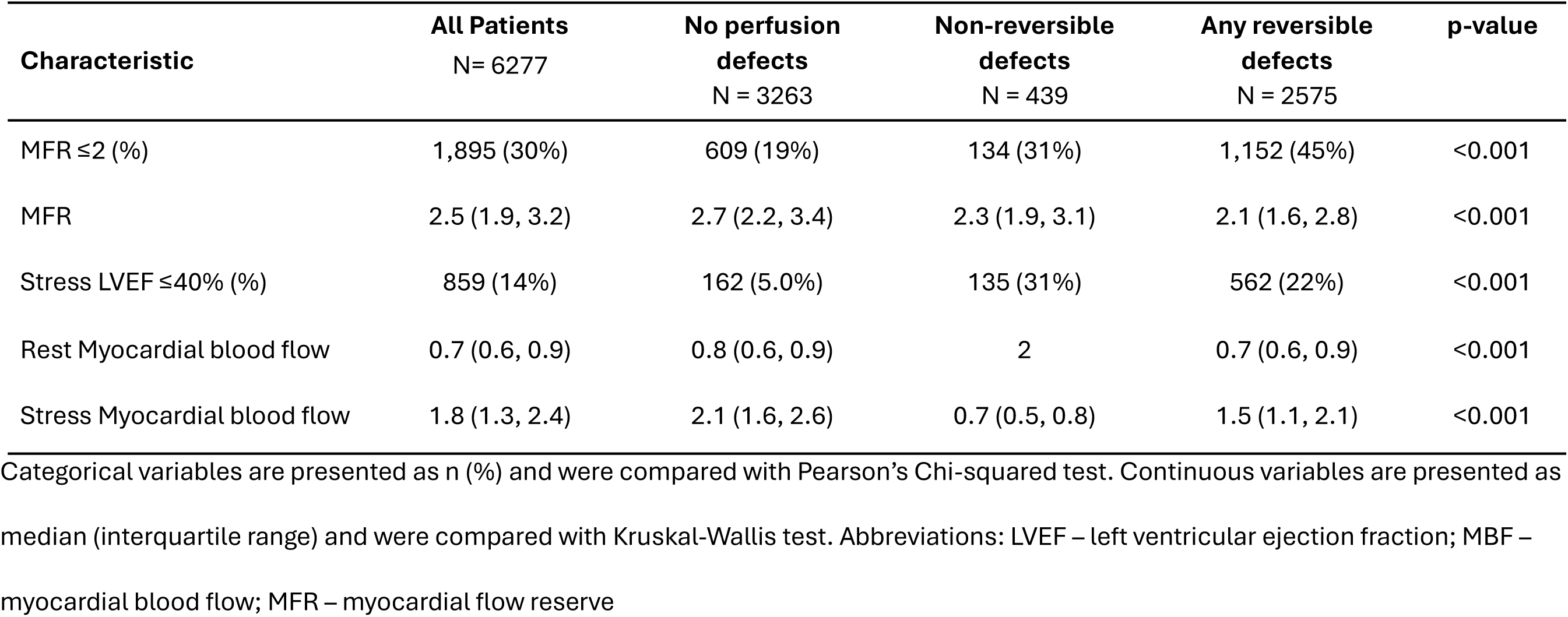
NH3 PET-Derived Variables Presented for Each Subgroup Defined by Extent and Type of Perfusion Defect.

| **Characteristic** | **All Patients**<br>**N= 6277** | **No perfusion**<br>**defects**<br>**N = 3263** | **Non-reversible**<br>**defects**<br>**N = 439** | **Any reversible**<br>**defects**<br>**N = 2575** | **p-value** |
| --- | --- | --- | --- | --- | --- |
| MFR ≤2 (%) | 1,895 (30%) | 609 (19%) | 134 (31%) | 1,152 (45%) | <0.001 |
| MFR | 2.5 (1.9, 3.2) | 2.7 (2.2, 3.4) | 2.3 (1.9, 3.1) | 2.1 (1.6, 2.8) | <0.001 |
| Stress LVEF ≤40% (%) | 859 (14%) | 162 (5.0%) | 135 (31%) | 562 (22%) | <0.001 |
| Rest Myocardial blood flow | 0.7 (0.6, 0.9) | 0.8 (0.6, 0.9) | 2 | 0.7 (0.6, 0.9) | <0.001 |
| Stress Myocardial blood flow | 1.8 (1.3, 2.4) | 2.1 (1.6, 2.6) | 0.7 (0.5, 0.8) | 1.5 (1.1, 2.1) | <0.001 |

Categorical variables are presented as n (%) and were compared with Pearson's Chi-squared test. Continuous variables are presented as median (interquartile range) and were compared with Kruskal-Wallis test. Abbreviations: LVEF – left ventricular ejection fraction; MBF – myocardial blood flow; MFR – myocardial flow reserve

### Clinical Outcome

During study follow-up of median 3.8 (IQR 2.2, 6.0) years, there were 1238 deaths. Mortality was significantly higher among patients with an MFR ≤2 (n=701, 37.0%) than patients with an MFR >2 (n=537, 12.3%; p< 0.001). Cause of death was available for 1033 (83.4%) of the patients. Among these, 443 (35.8%) deaths were attributed to cardiovascular causes.

### MFR Deciles and Associated Risk of Mortality

**Supplementary Figure 1A** presents unadjusted hazard ratios of ACM across MFR deciles in the total study cohort. Lower MFR values (deciles 1–3) are associated with a significantly increased risk of ACM, whereas higher MFR values (deciles 5–10) correspond to a reduced risk. Decile 4, which includes participants with MFR values between 1.99 to 2.23, showed no significant difference in mortality compared to the rest of the population (HR 0.85, 0.70 – 1.04; p=0.111). However, results for Decile 4 improved after adjustment for age and sex (HR 0.73, 95% CI 0.60–0.89; p=0.002) (**Figure 1A**). Detailed adjusted and unadjusted HRs per MFR decile are in **Supplementary Table 2**.

**Figure 1.**
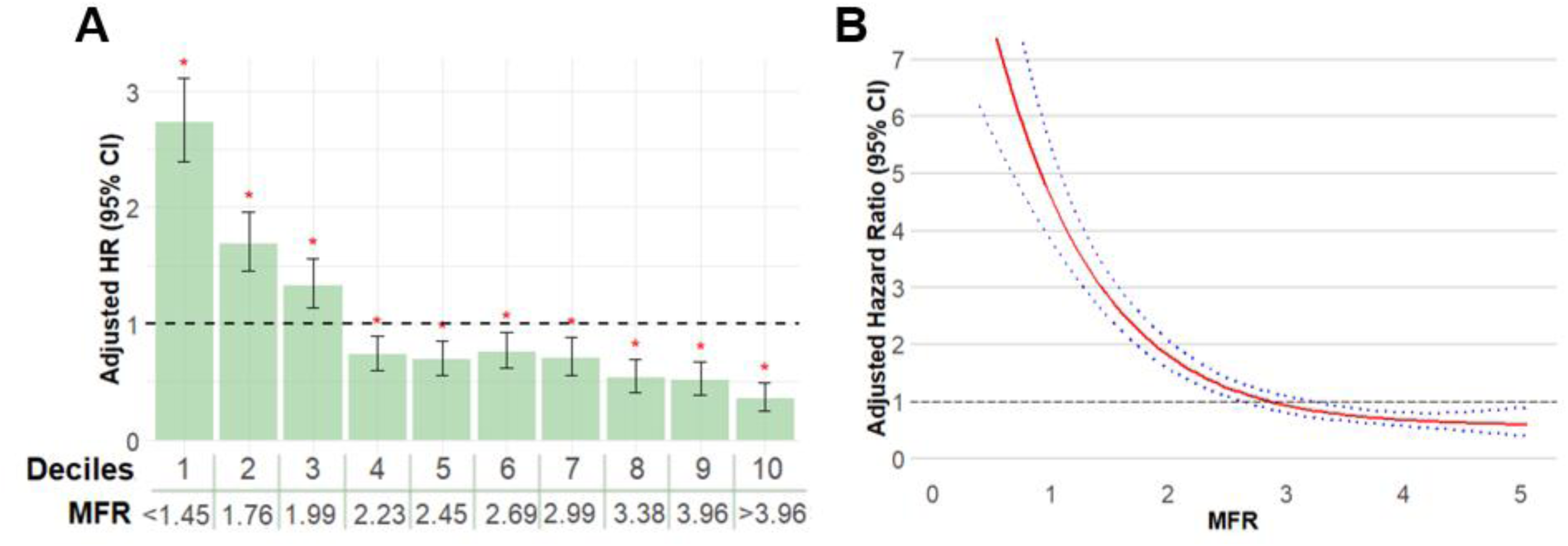
(A) Adjusted HRs of ACM by MFR Deciles and (B) Continuous HR Analysis of ACM. HRs were adjusted for age and sex. (A) Each bar represents a decile (10% interval) of MFR with 95% confidence intervals around the HR estimates. The x-axis shows the MFR threshold for each decile group. Lower MFR (deciles 1–3) is associated with an increased risk for all-cause mortality, whereas higher MFR (deciles 4–10) is associated with a decreased risk. The dashed horizontal line represents the reference value. Red asterisks indicate a statistically significant difference as compared to the reference. (B) Estimated HRs (red solid line) and 95% CI’s (blue dashed line). Abbreviations: CI – confidence interval; HR – hazard ratio; MFR - myocardial flow reserve

### Continuous MFR Analysis for Risk of Mortality

When evaluating hazard ratios on a continuous scale, MFR values from 2.41 - 2.45 were associated with lowest change in risk for ACM in unadjusted analysis (HR 1.02, 0.96 – 1.08; p=0.496 to HR 0.98, 0.92 – 1.04; p=0.46) (**Supplementary Figure 1B**). In the adjusted analysis accounting for age and sex (**Figure 1B**), the neutral range shifted slightly higher numerically, with MFR values from 2.78 - 2.83 showing lowest change in mortality risk (HR 1.01, 0.87 – 1.16; p=0.937 to HR 0.98, 0.84 – 1.14; p=0.796).

### MFR and Risk of ACM

Overall, survival rates declined as MFR decreased (**Figure 2**). Unadjusted and adjusted hazard ratios with adjustments to sex and age, in reference to MFR>2.5 group, demonstrate statistical significance.

**Figure 2.**
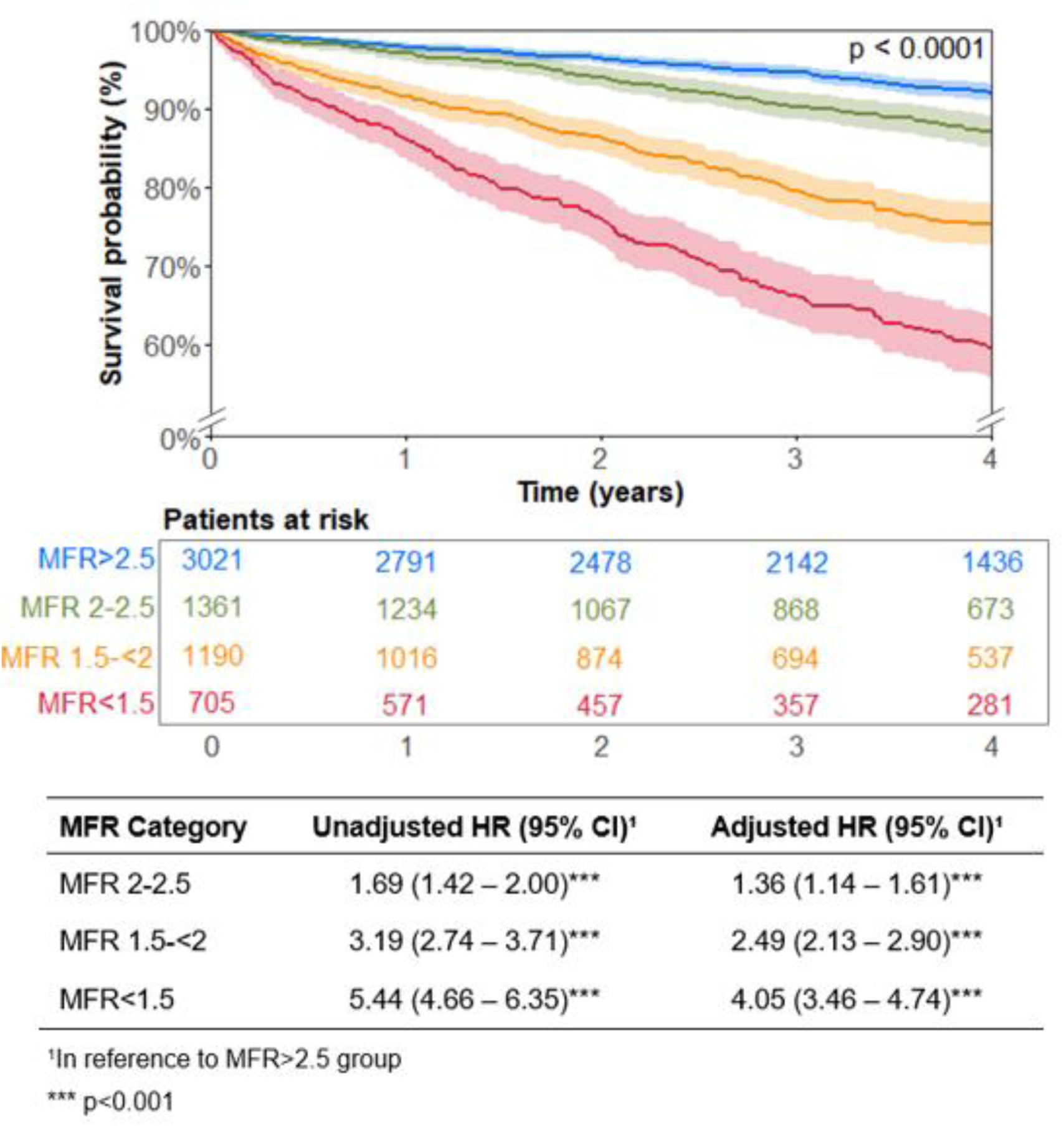
Survival curves for All-cause Mortality Stratified by Myocardial Flow Reserve (MFR). Patients with MFR <1.5 (red) had highest risk of all-cause mortality over 4 years, followed by those with MFR 1.5-<2 (orange), MFR 2-2.5 (green), while patients with MFR >2.5 (blue) had the lowest risk. Number of overall population size at time zero is given by “n”. Numbers boxed below the curve represent the number of patients at risk at each time point. P-values were calculated by the Log-rank test.

Patients with MFR ≤2 had a significantly lower overall survival than those with MFR >2 regardless of the extent of reversible or non-reversible perfusion defects (**Figure 3**). MFR ≤2 remained a strong independent predictor of ACM among patients with no perfusion defects (unadjusted HR 2.88, 95% CI 2.36-3.51; p<0.001), including when adjusted for clinical and imaging covariates as shown in **Table 3**. Even with patients with a previous history of coronary artery disease or not, displayed a similar trend where patients with MFR≤2 were at a higher risk of ACM than patients with MFR<2 (unadjusted HR 2.80 [95% CI 2.35 – 3.32] and 3.09 [95% CI 2.65 – 3.60], respectively; p<0.001 for all) in **Supplementary Figure 2**.

**Figure 3.**
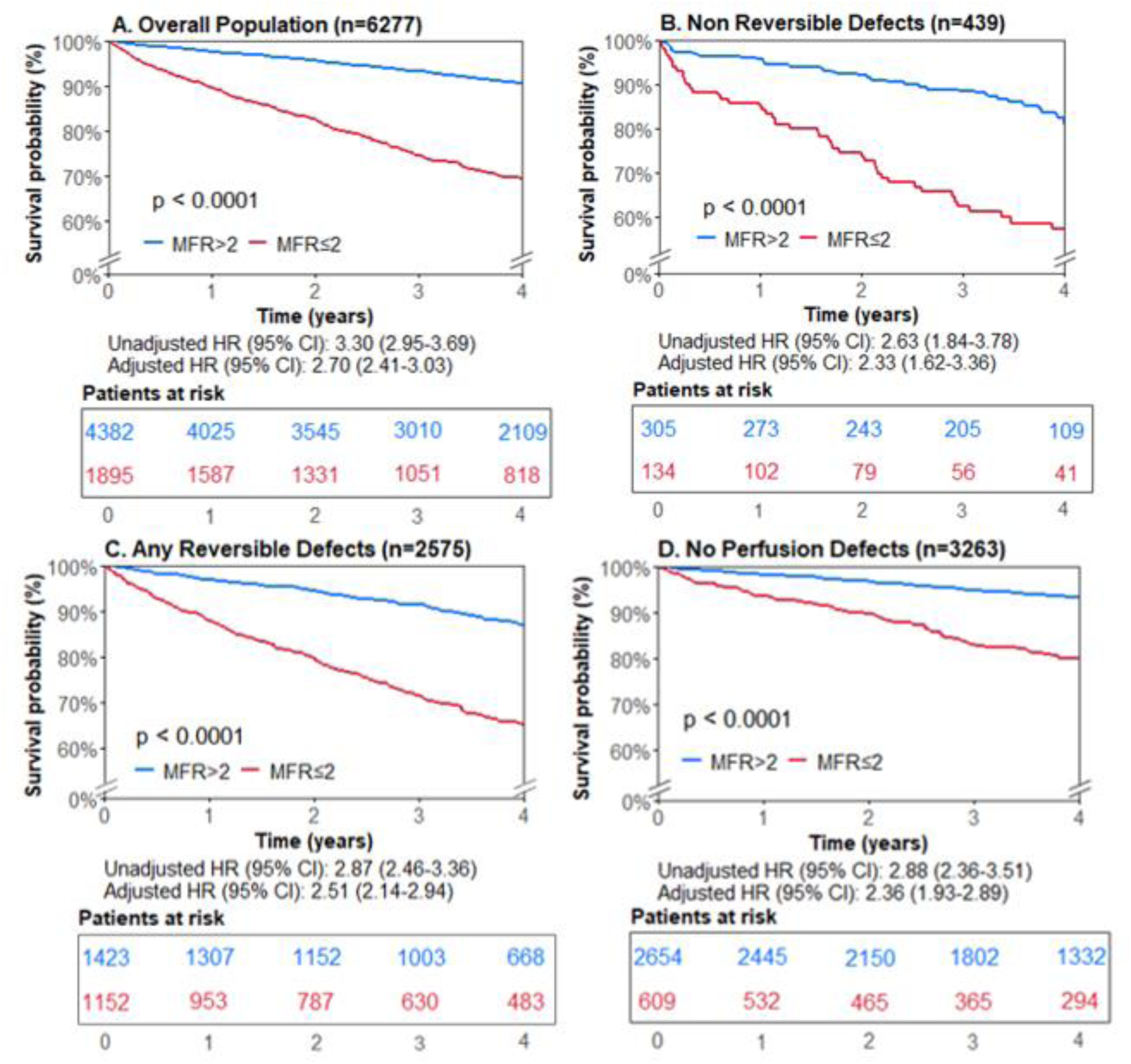
Survival Curves for All-cause Mortality Stratified by Myocardial Flow Reserve (MFR) and Perfusion Defects. Survival curves stratified by MFR in the overall population (A) and subgroups (B-D) defined by the extent and type of perfusion defects. MFR >2 is presented in blue and MFR ≤2 in red. Hazard ratios were adjusted for age and sex. Number of population size at time zero are given by “n”. Numbers boxed below each curve represent the number of patients at risk at each time point. P-values were calculated by the Log-rank test. Abbreviations: HR – hazard ratio; CI – confidence interval

**Table 3.** Cox Survival Regression Model of All-Cause Mortality in 3263 Patients with No Perfusion Defects.

| Variable | Crude Model HR<br>(95% CI) | Model 1<br>HR (95% CI) | Model 2<br>HR (95% CI) |
| --- | --- | --- | --- |
| MFR ≤2 | 2.88 (2.36-3.51)*** | 2.33 (1.90-2.85)*** | 2.16 (1.76-2.69)*** |
| Age 60-70 | - | 1.61 (1.22-2.12)*** | 1.50 (1.13-1.98)** |
| Age 70-80 | - | 2.94 (2.24-3.86)*** | 2.59 (1.96-3.42)*** |
| Age ≥80 | - | 5.41 (3.95-7.42)*** | 4.48 (3.23-6.21)*** |
| Sex (male) | - | 1.48 (1.21-1.80)*** | 1.37 (1.12-1.67)** |
| Previous CAD | - | 1.26 (0.99-1.61) | 1.26 (0.99-1.60) |
| BMI | - | - | 0.98 (0.97-0.99)** |
| Heart Rate (10bpm Increase) | - | - | 0.60 (0.48-0.74)*** |
| LVEF (10% Increase) | - | - | 0.71 (0.34-1.51) |

Significance: \*\* p<0.01, \*\*\* p<0.001
Model 1 was adjusted for MFR, age (Age<60 as reference group), sex, and prior CAD (defined as history of myocardial infarction or revascularization). Model 2 was additionally adjusted for BMI, heart rate, and LVEF. Abbreviations: BMI – body mass index; bpm – beats per minute; CAD – coronary artery disease; CI – confidence interval; HR – hazard ratio; LVEF – left ventricular ejection fraction

### Rates of ACM and Cardiovascular Mortality

Among the 26,708 total person-years of follow-up, the cardiovascular mortality cases were over threefold higher in the MFR ≤2 group (3.6 vs. 0.9 per 100 person-years), as was non-cardiovascular mortality (4.3 vs. 1.4 per 100 person-years) (**Table 4**). Similarly, patients with MFR ≤2 had higher observed incidence rates of ACM compared to those with preserved MFR (9.3 vs 2.8 deaths per 100 person-years, respectively). Annualized ACM rates ranged from 1.7% to 11.6% and cardiovascular mortality rates ranged from 0.5% to 4.9% depending on the combined SSS and MFR group (**Figure 4 and Supplementary Figure 3, respectively**). The annual rates of both ACM and cardiovascular mortality increased with increasing SSS and decreasing MFR; however, lower MFR consistently identified higher risk patients at every level of SSS, including among those with visually normal scans.

**Figure 4.**
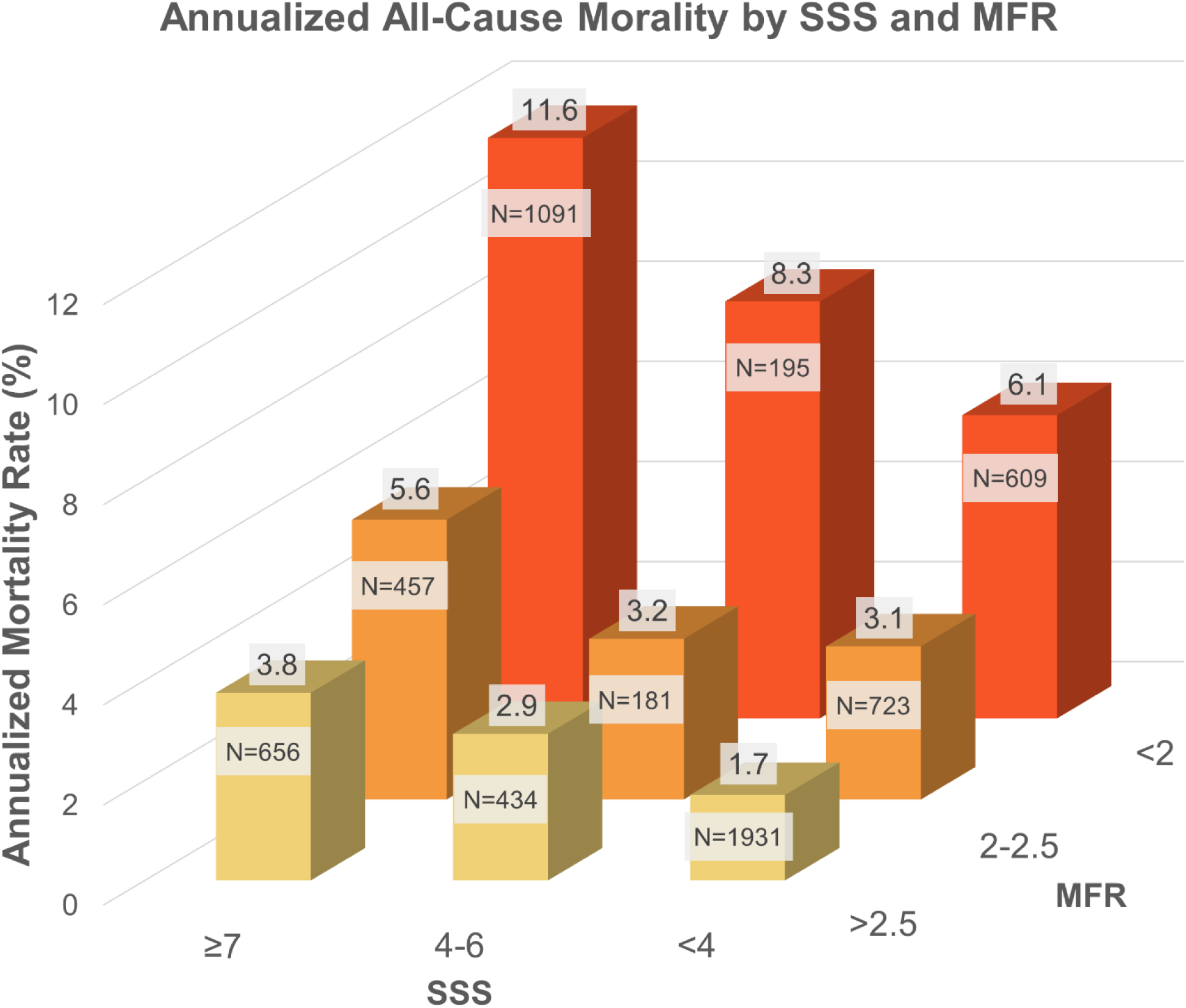
Annualized All-Cause Mortality Rates by SSS and MFR. Each bar represents the annualized mortality rate per 100 person years for that group. The x-axis represents MFR groups, the y-axis represents SSS groups, and the z axis the annualized mortality rate (%). Numbers above each bar indicate the annualized mortality rate for that group. Number of total patients in each group given by “N”. Abbreviations: SSS – summed stress score; MFR – myocardial flow reserve

**Table 4.** Annualized Mortality Rates Stratified by MFR.

| | Overall | MFR≤2 | MFR>2 |
| --- | --- | --- | --- |
| All-cause mortality | 4.6 | 9.3 | 2.8 |
| Person-years | 26,708 | 7,552 | 19,156 |
| Cause of death known | 3.9 | 7.9 | 2.3 |
| Cardiovascular death | 1.7 | 3.6 | 0.9 |

Annualized mortality rates per 100 person-years for all-cause mortality, cardiovascular death and non-cardiovascular death. Cause of death is known when occurred between 0 to 4645 days (13 years) after PET and was available for n=1,033 (83.4% of all deaths). Of these patients, 270 had cardiovascular death with MFR ≤ 2 and 173 had cardiovascular death with MFR >2. Abbreviations:
CI – confidence interval; MFR – myocardial flow reserve

The relationship between MFR and global stress MBF, along with the distribution of all-cause and cardiovascular deaths, is illustrated in the scatter plots shown in **Figure 5**. The highest concentration of all-cause and cardiovascular deaths was observed among patients with concordant MFR and MBF impairment. Among patients with discordant findings, those with impaired MFR but preserved MBF had a higher annualized rate of mortality (8.0 for ACM and 2.2 for cardiovascular death) than those with preserved MFR and impaired MBF (3.4 and 1.2, respectively).

**Figure 5.**
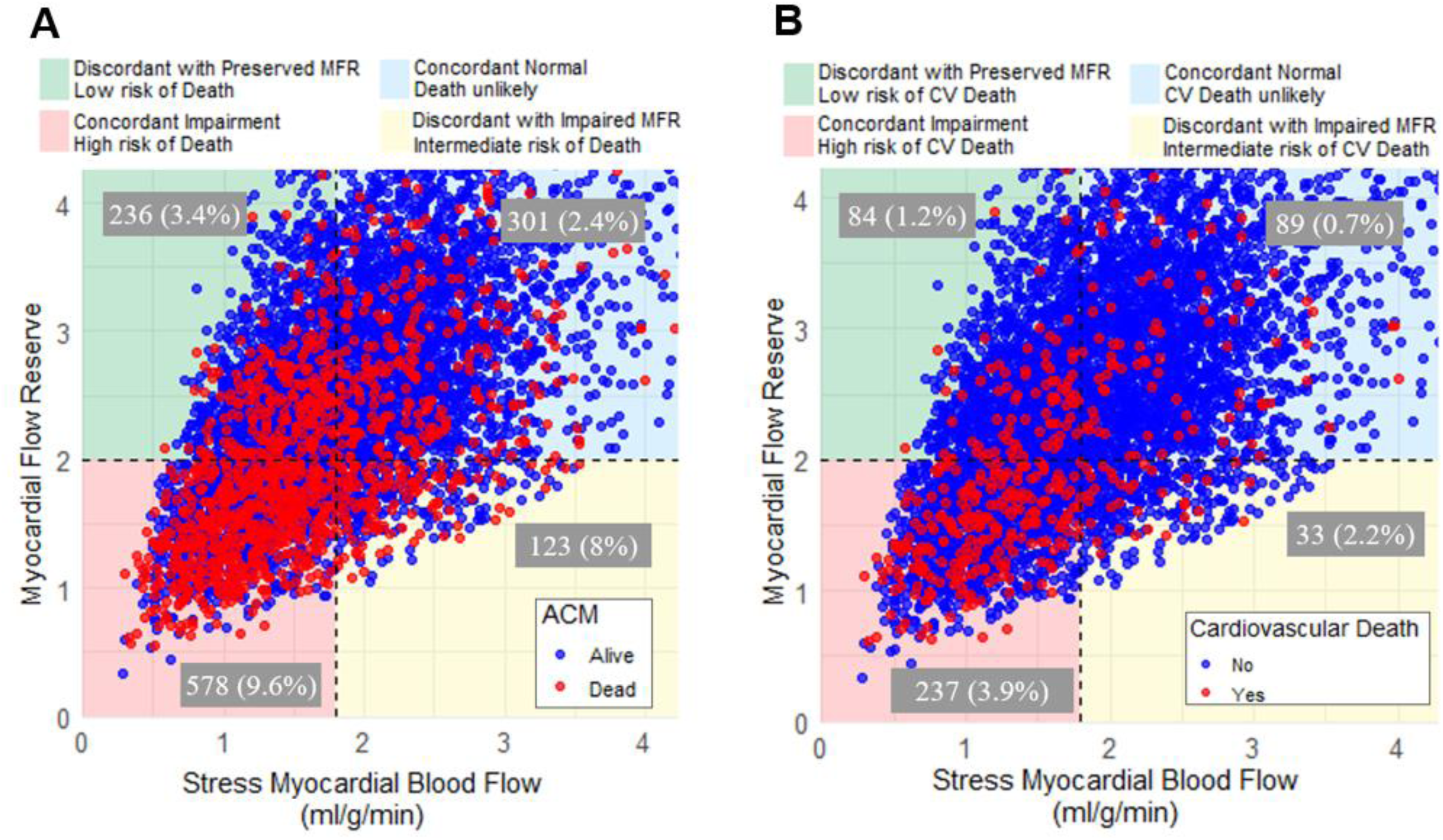
Scatter plot of MFR and Global MBF by Annualized (A) ACM and (B) Cardiovascular Death Event Rates. MFR <2 and global MBF <1.8 ml/g/min were defined as impaired. Abbreviations: MFR – myocardial flow reserve; MBF – myocardial blood flow

## DISCUSSION

In this multicenter study of 6277 patients undergoing ^13^N-ammonia PET MPI at five sites across the United States and Mexico, MFR demonstrated strong and independent prognostic value for ACM. Importantly, this relationship held regardless of the presence or absence of perfusion defects— reversible or fixed—and remained significant after adjustment for traditional cardiovascular risk factors including age, sex, prior CAD, BMI, heart rate, and LVEF. Patients with impaired MFR (≤2.0) had more than a threefold higher risk of mortality compared to those with preserved MFR (>2.0). Notably, MFR consistently identified higher-risk individuals across the spectrum of SSS, suggesting its value as a complementary marker of physiologic risk.

Beyond perfusion findings, MFR with ^13^N-ammonia PET provided incremental prognostic information relative to both TPD and absolute MBF. In discordant cases, individuals with impaired MFR but preserved MBF experienced higher rates of both ACM and cardiovascular mortality than those with preserved MFR and impaired MBF—highlighting ^13^N-ammonia PET MFR’s unique ability to detect latent physiologic abnormalities not captured by flow alone.

This study builds upon a growing body of evidence demonstrating the prognostic utility of MFR measured by PET MPI (3). While previous large-scale investigations have primarily employed ^82^Rubidium PET (17-21), ^15^O-water PET (22, 23), or mixed-tracer (24, 25), studies specifically using ^13^N-ammonia PET have been comparatively fewer and generally limited to smaller cohorts (26-28), post-revascularization populations(29), or specific subgroups such as patients with cardiomyopathy(30) or sex-stratified analyses(31). Other investigations have explored MFR’s associations with alternative imaging markers—including myocardial strain(32), CAD-RADS classification(32), and LVEF(33)—but not the direct relationship of ^13^N-ammonia PET-derived MFR with mortality outcomes in general populations.

Our findings extend prior work by evaluating ^13^N-ammonia PET–derived MFR in a large, diverse cohort and linking it to hard clinical endpoints. Using standardized, commercially available software across multiple sites, we assessed MFR’s prognostic impact across the full spectrum of perfusion patterns and stratified risk by decile. This allows for a more nuanced understanding of MFR’s clinical relevance, particularly in patients without overt perfusion defects. Importantly, we demonstrated that MFR retains prognostic significance even in patients with visually normal perfusion. In routine clinical practice, such patients are often considered low risk; however, our findings—and those of other recent studies (5)—suggest that impaired MFR in this group may reflect underlying endothelial dysfunction, diffuse atherosclerosis, or microvascular disease. These results support the role of ^13^N-ammonia PET–derived MFR in enhancing risk stratification for patients with suspected non-obstructive CAD, although further prospective study is warranted in this population.

We also identified a physiologically meaningful threshold for impaired MFR. In our cohort, an MFR around 2.23 represented a statistical inflection point for mortality risk (**Figure 1 and Supplementary Figure 1**). Definitions of impaired MFR vary widely in the literature, ranging from 1.49 to 2.4, but our results reinforce the validity of the most commonly used clinical threshold of ≤2.0 (3) as a meaningful cutoff for identifying increased risk in ^13^N-ammonia PET MPI. Furthermore, risk declined progressively with increasing MFR, both across deciles and on a continuous scale.

Patients in the highest MFR percentiles exhibited the lowest hazard ratios for mortality, suggesting that high MFR may serve not only as a marker of cardiovascular reserve but potentially of overall physiologic resilience and wellness.

### Study Limitations

Our study has several limitations. First, global MFR rather than regional MFR was considered in the present analysis. Global MFR reflects an average of flow across the entire myocardium and thus limits the ability to detect localized impairments. However, global MFR effectively captures both severity and extent of perfusion abnormalities and is especially valuable for assessing microvascular dysfunction. Future research should focus on evaluating the prognostic utility of regional MFR abnormalities and their incremental value beyond global MFR assessment. Our cohort consisted of patients referred for PET MPI at quaternary care centers due to known or suspected CAD, representing a relatively high-risk population. This referral bias may amplify the observed associations between MFR and mortality. Further evaluation in lower-risk, diverse patient populations is needed. Data for cardiac versus non-cardiac death were not available for all study participants. We were able to perform this analysis on 83.4% of deceased participants. Finally, resting MBF was not corrected for the rate-pressure product (RPP), which may contribute to higher resting MBF estimates. However, it has previously been reported that impaired MFR is associated with increased risk of MACE regardless of RPP correction (34).

### Conclusions

MFR assessed with ¹³N-ammonia PET MPI is a powerful and independent predictor of ACM, including in patients with normal relative perfusion. An MFR threshold of ≤2.0 was associated with a more than threefold increase in mortality risk, while progressively higher MFR values correlated with improved survival, underscoring its role as a marker of cardiovascular health and physiologic resilience. These findings reinforce the clinical utility of MFR and support its routine incorporation into ¹³N-ammonia PET MPI to enhance risk stratification and inform the management of patients with suspected or known CAD.

## Data Availability

To the extent allowed by data sharing agreements and IRB protocols, the deidentified data and data analysis code from this manuscript will be shared upon written request.

## Abbreviations

ACM: All-Cause Mortality
BMI: Body Mass Index
CAD: Coronary Artery Disease
MBF: Myocardial Blood Flow
MFR: Myocardial Flow Reserve
MI: Myocardial Infarction
MPI: Myocardial Perfusion Imaging
PET: Positron Emission Tomography
REFINE PET: REgistry of Fast Myocardial Perfusion Imaging with NExt generation PET
RPP: Rate-Pressure Product
SDS: Summed Difference Score
SRS: Summed Rest Score
SSS: Summed Stress Score
TPD: Total Perfusion Deficit

## Supplemental Material

Supplementary Tables 1–2

Supplementary Figures 1-3

