## Supplemental Material for "Multicenter Evaluation of Myocardial Flow Reserve as a Prognostic Marker for Mortality in ¹³N-Ammonia PET Myocardial Perfusion Imaging"

**SUPPLEMENTARY MATERIAL**

### Supplementary Tables

***Supplementary Table 1. Missingness in Demographic/Clinical Data***

| Demographic/Clinical Variable | Missingness N (%) |
|---|---|
| Race | 673 (10.72) |
| BMI | 7 (0.11) |
| Hypertension | 29 (0.46) |
| Dyslipidemia | 29 (0.46) |
| Diabetes | 1 (0.02) |
| Smoking | 28 (0.45) |
| Prior CAD | 48 (0.76) |
| Past PCI/stents | 48 (0.76) |
| Past MI | 48 (0.76) |
| Past Bypass | 48 (0.76) |
| Stress LVEF | 50 (0.80) |
| Rest LVEF | 60 (0.96) |
| Resting BP Systole | 46 (0.73) |
| Stress Systolic BP-Peak | 35 (0.56) |
| Resting Heart Rate | 1 (0.02) |
| Stress Heart Rate-Peak | 3 (0.05) |

Abbreviations: BMI – body mass index; CAD – coronary artery disease; PCI – percutaneous coronary intervention; MI – myocardial infarction; PVD – peripheral vascular disease; LVEF – left ventricular ejection fraction; BP – blood pressure

***Supplementary Table 2. Hazard Ratios (HR) for Mortality*** **by Myocardial Flow Reserve (MFR) Deciles**

| MFR Percentile Groups | Values | Unadjusted HR (95% CI) | p-value | Adjusted HR* (95% CI) | p-value |
|---|---|---|---|---|---|
| 0 - <10 | <1.45 | 3.35 (2.94 - 3.81) | <0.001 | 2.73 (2.39 - 3.11) | <0.001 |
| 10 - <20 | 1.45-<1.76 | 2.05 (1.77 - 2.38) | <0.001 | 1.69 (1.45 - 1.96) | <0.001 |
| 20 - <30 | 1.76-<1.99 | 1.50 (1.28 - 1.76) | <0.001 | 1.33 (1.14 - 1.56) | <0.001 |
| 30- <40 | 1.99-<2.23 | 0.85 (0.70 - 1.04) | 0.111 | 0.73 (0.60 - 0.89) | 0.002 |
| 40 - <50 | 2.23-<2.45 | 0.76 (0.62 - 0.93) | 0.008 | 0.70 (0.57 - 0.86) | <0.001 |
| 50 - <60 | 2.45-<2.69 | 0.79 (0.64 - 0.96) | 0.021 | 0.75 (0.61 - 0.92) | 0.006 |
| 60 - <70 | 2.69-<2.99 | 0.62 (0.50 - 0.78) | <0.001 | 0.70 (0.56 - 0.88) | 0.002 |
| 70 - <80 | 2.99-<3.38 | 0.47 (0.36 - 0.6) | <0.001 | 0.53 (0.41 - 0.69) | <0.001 |
| 80 - <90 | 3.38-<3.96 | 0.39 (0.30 - 0.52) | <0.001 | 0.51 (0.39 - 0.67) | <0.001 |
| 90 - <100 | ≥3.96 | 0.25 (0.18 - 0.35) | <0.001 | 0.35 (0.25 - 0.50) | <0.001 |

Reference group is all other patients not in that specified MFR percentile group.

Abbreviations: CI – confidence

*Adjusted for age and sex.

**Supplementary Figures**

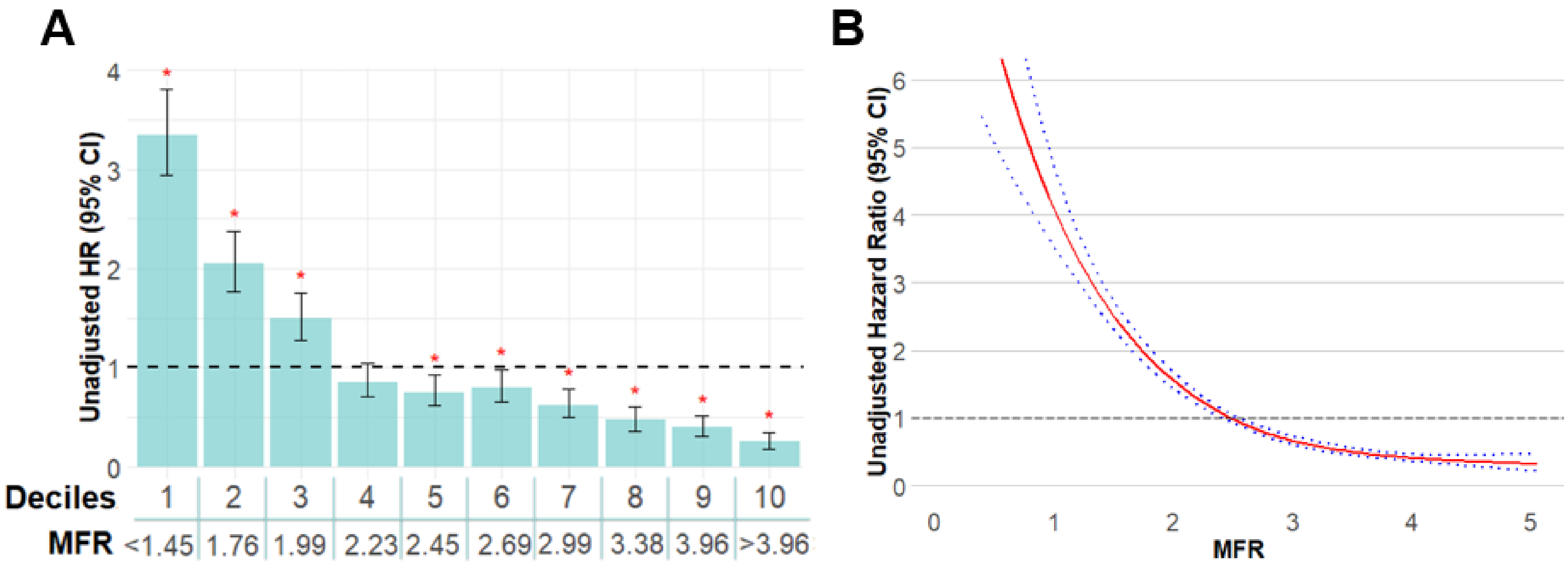


**Supplementary Figure 1. (A) Unadjusted HRs of ACM by MFR Deciles and (B) Continuous HR Analysis of ACM.** (A) Each bar represents a decile (10% interval) of MFR with 95% confidence intervals around the HR estimates. The x-axis shows the MFR threshold for each decile group. Lower MFR (deciles 1–3) is associated with an increased risk for all-cause mortality, whereas higher MFR (deciles 5–10) is associated with a decreased risk. The dashed horizontal line represents the reference value. Red asterisks indicate a statistically significant difference as compared to the reference. (B) Estimated HRs (red solid line) and 95% CI's (blue dashed line). Abbreviations: CI – confidence interval; HR – hazard ratio; MFR - myocardial flow reserve.

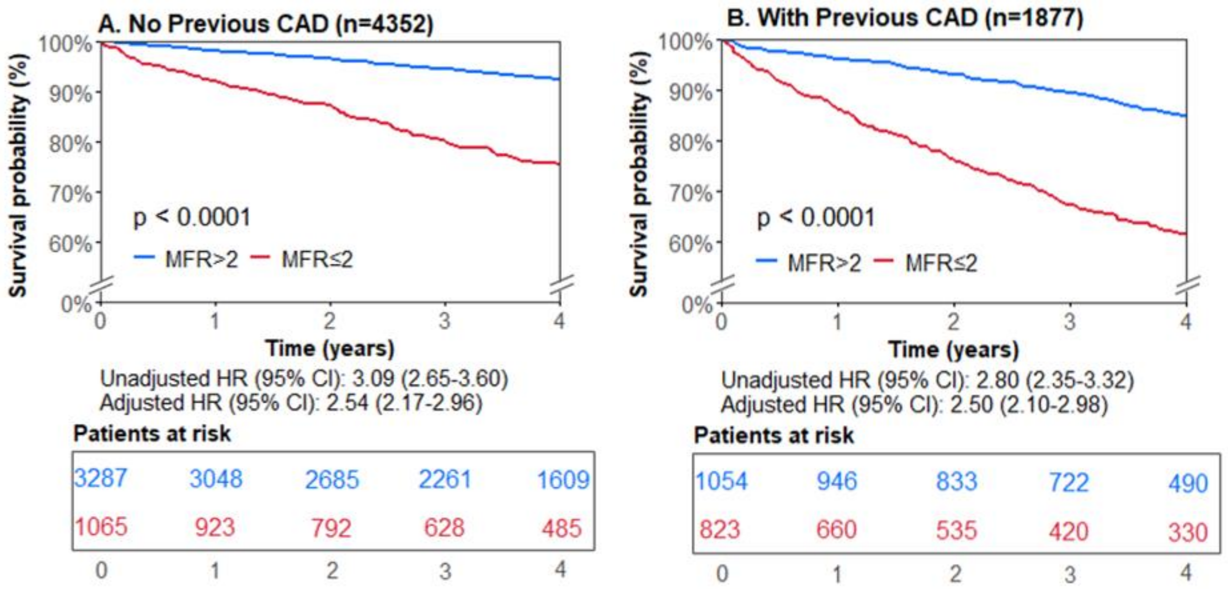


**Supplementary Figure 2. Survival Curves for All-cause Mortality Stratified by Myocardial Flow Reserve (MFR) and Previous CAD.** Survival curves stratified by MFR in subgroups (A-B) by history of CAD. MFR >2 is presented in blue and MFR ≤2 in red. Hazard ratios were adjusted for age and sex. Number of population size at time zero are given by "n". Numbers boxed below each curve represent the number of patients at risk at each time point. P-values were calculated by the Log-rank test.

Abbreviations: CAD – coronary artery disease; HR – hazard ratio; CI – confidence interval

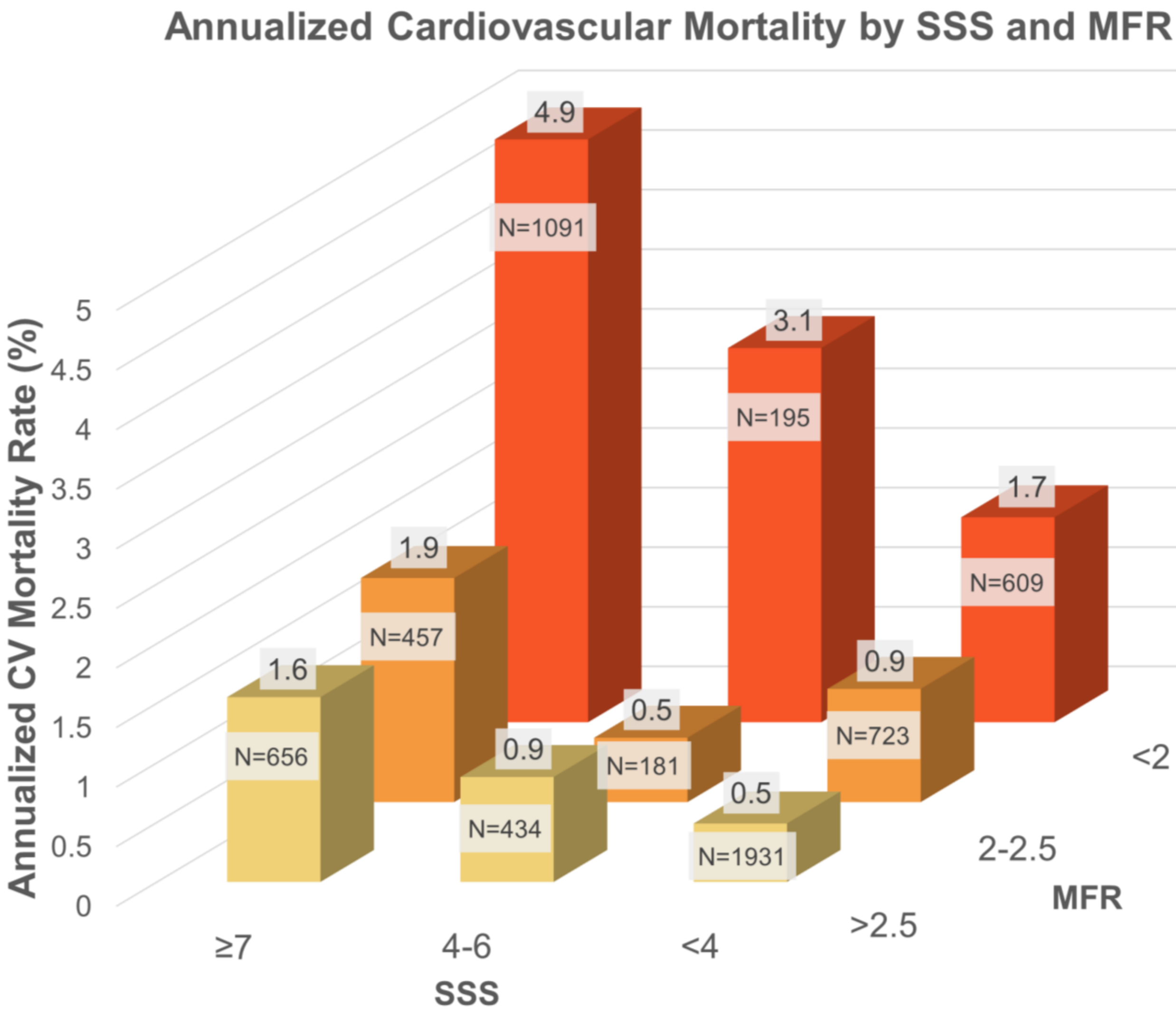


**Supplementary Figure 3. Annualized Cardiovascular Mortality Rates by SSS and MFR.** Each bar represents the annualized cardiovascular mortality rate for that group. The x-axis represents MFR groups, the y-axis represents SSS groups, and the z axis the annualized mortality rate (%). Numbers above each bar indicate the annualized mortality rate for that group. Number of total patients in each group given by "N". Abbreviations: CV – cardiovascular; SSS – summed stress score; MFR – myocardial flow reserve
